# Drinking habits and executive functioning: a propensity score-weighted analysis of 78,832 adults

**DOI:** 10.1101/2020.12.22.20248655

**Authors:** Lizanne JS Schweren, Jan Haavik, Lin Li, Berit Skretting Solberg, Henrik Larsson, Catharina A Hartman

## Abstract

Excessive alcohol intake compromises cognitive functioning. At the same time, moderate alcohol consumption is reported to protect against Alzheimer’s disease among elderly. Little is known about dose-dependent effects of alcohol consumption on higher-order cognitive functioning among generally healthy adults. Here, we applied propensity weighted analyses to investigate associations between habitual drinking patterns and executive functioning in the general population.

A community sample of N=78,832 Dutch adults (age 18-65, 40.9% male) completed the Ruff Figural Fluency Task of executive functioning (range 1-165), and self-reported their past month consumption of alcoholic beverages on a food frequency questionnaire. Participants were stratified according to drinking level (abstinent [22.0%], occasional [<2.5 g/day, 21.4%], light [2.5–14.9 g/day, 42.9%], moderate [15–29.9 g/day, 11.4%], or heavy [>30 g/day, 2.3%]) and binge-drinking (yes [10.6%] vs. no [89.4%]). Groups were equivalised using multinomial propensity score weighing based on demographic, socioeconomic, health-related and psychosocial factors influencing drinking behaviour.

Compared to abstinent participants, task performance was better among light drinkers (β[95% CI]=0.056[0.033-0.078] or +1.3 points, p<0.0001) and moderate drinkers (β[95% CI]=0.111[0.079-0.143] or +2.5 points, p<0.0001), but not among occasional drinkers (β[95% CI]=0.018[-0.006-0.043], p=0.1432) or heavy drinkers (β[95% CI]=0.075[-0.009-0.158], p=0.0791). No difference was found between binge-drinkers and non-binge-drinkers (β[95% CI]=0.032[-0.002-0.066], p=0.0654).

In conclusion, we observed better executive functioning in light-moderate drinkers compared to abstainers, but not in occasional drinkers, heavy drinkers or binge-drinkers. This non-linear association is reminiscent of the dose-dependent effect of alcohol consumption on cardiovascular risk. Further studies may determine whether cardiovascular, inflammatory and/or other somatic factors mediate the association between moderate drinking and higher-order cognitive functions. Although analyses were adjusted for observed factors that influence drinking behaviours, we cannot exclude a contribution from unobserved residual confounding.

## Introduction

Recent years have witnessed a shift in alcohol consumption guidelines worldwide. Instigated by the World Health Organisation [1], several national health institutes (e.g. UK [2]) now proclaim that there is no safe level of alcohol consumption. The same departure from prior guidelines, in which one respectively two drinks per day was deemed safe for adult males and females is expected to take effect in the United States in 2020 [3].

While for certain lifestyle behaviours (e.g. smoking) the continuity hypothesis of dose-dependent effects is well established, this assumption is heavily debated with respect to drinking outcomes. Low levels of alcohol consumption increase the risk of certain negative outcomes (e.g. accident-related injuries [4]), but not for others. For instance, for cardiovascular outcomes, both abstinence and very heavy drinking are associated with higher risks compared to light of moderate drinking [5], [6]. Such effects might be mediated by the blood pressure-lowering properties of alcohol [7], and/or by anti-inflammatory or anti-oxidative processes [8].

Little is known about dose-dependent effects of alcohol intake on higher-order cognitive functioning. Executive functions refer to a set of cognitive processes necessary for flexible and goal-directed behaviours, including working memory, planning, inhibitory control, self-monitoring and problem solving [9]. Executive functioning shows moderate genetic overlap with disinhibited psychiatric phenotypes including attention-deficit/hyperactivity disorder and alcohol dependence, but much less so with continuous measures of (non-problematic) alcohol intake [10]. Associations between drinking and executive functioning have predominantly been described in three groups that are not representative of the general population. First, abundant evidence shows that patients with a diagnosis of alcohol use disorder exhibit deficits on a range of executive tasks [11]. Second, among adolescents and students, heavy social drinking or binge-drinking is typically not found to result in poorer executive functioning [12]–[16]. It has been suggested that compensatory mechanisms may dissipate negative effects of repeated alcohol intoxication on the developing brain [17], or that detrimental effects may appear at later age as a result of cumulative exposure [18]. The third group concerns elderly at increased risk for cognitive decline. Adjusting for socio-economic confounders, moderate drinking is associated with better executive functioning in this group [19]–[21], but does not protect against age-related cognitive decline over time [20]–[23].

The above associations cannot readily be generalised to high-functioning, non-student and non-elderly populations. Yet, very few studies have quantified the association between different levels of alcohol consumption and higher-order cognitive functioning among generally healthy adults. Rodgers compared working memory performance between abstainers, occasional drinkers, light-, moderate- and harmful drinkers among 7,485 participants age 20-24, 40-44 or 60-64. Analyses, in which age groups were collapsed, revealed better task performance among light drinkers (up to 20/10 grams per day in men/women, respectively) compared to abstainers or occasional drinkers [24]. By contrast, among 3,361 males age 31-49, neither verbal fluency nor abstract reasoning was associated with years of drinking or drinking intensity [25]. Finally, among 280 couples (average age 36) oversampled for heavy drinking, number of drinks per day was negatively associated with task performance on executive tasks tapping planning, inhibition and set-shifting [26]. However, the latter two studies did not model non-linear associations, and that no study included other health-related behaviours (e.g. physical activity) or income as potential confounders.

Here, in a community-sample of almost 80,000 Dutch adults aged 18 to 65 years old, we aimed to investigate associations between habitual alcohol consumption and figural fluency task performance as an indicator of executive functioning. We modelled linear and non-linear effects of multiple drinking levels (abstinent, occasional, light, moderate, heavy) and patterns (binge-drinking vs. non-binge-drinking), applying propensity weighted analyses to address non-random group allocation.

## Methods

### Data source and eligibility

The Lifelines Cohort Study is a multi-disciplinary prospective population-based cohort study examining the health of 167,729 persons living in the North of the Netherlands [27]. Baseline data regarding biomedical, socio-demographic, behavioural and psychological factors were collected between 2006 and 2015. Data collection involved self-administered questionnaires, in-person interviews, blood sample and DNA collection, and physiological measurements at one of the Lifelines research sites. Written informed consent was obtained from all participants. The Lifelines Cohort Study is conducted according to the principles of the Declaration of Helsinki and is approved by the medical ethics committee of the University Medical Center Groningen, The Netherlands.

Randomly selected general practitioners in three northern provinces of the Netherlands invited all their listed patients between 25 and 50 years of age. Almost all inhabitants of the Netherlands are registered with a local general practitioner. Willing respondents and their family members were asked to participate. Subjects could also register themselves at the Lifelines website. Exclusion criteria for the Lifelines study were: a) severe mental or physical illness, b) unable to visit the general practitioner, c) unable to complete the questionnaires, and d) insufficient understanding of the Dutch language. For the current study, baseline data were used applying the following additional exclusion criteria: e) age <18 or >65 years; f) missing or incomplete data on executive functioning or drinking habits (see below); g) self-reported diagnosis of any neurological disorder or dementia at baseline; and h) for participants aged 65, Mini Mental State Examination (MMSE)-score below 26. This resulted in a final sample of N=78,832 participants with an average age of 42.8 (SD=10.8) years of whom N=32,267 (40.9%) male (see flowchart, Supplement 1).

### Exposure: habitual alcohol intake

Past-month food and drink intake was assessed using a 110-item semi-quantitative food frequency questionnaire [28]. Participants self-reported a) past month consumption of alcoholic drinks on a seven-point ordinal scale ranging from “never/not this month” to “6–7 days a week”; b) average number of glasses per day on such occasions; and c) how often (never/sometimes/often/always) on such occasions they drank beer, red or rose wine, white wine, fortified wine (e.g. sherry, port), liquor/distilled alcoholic drinks (e.g. rum, whiskey), and other alcoholic drinks. Items were weighted by their alcohol content (one Dutch standard unit [SU_NL_] = 10 grams of alcohol) and summed across drink types to derive alcohol intake in grams per day.

Two indicators of habitual alcohol intake were defined. First, based on a meta-analysis regarding alcohol intake and cardiovascular risk [5], drinking level was categorised as abstinent, occasional (<2.5 g/day or <0.25 SU_NL_/day), light (2.5–14.9 g/day or 0.25-1.5 SU_NL_/day), moderate (15–29.9 g/day or 1.5-3 SU_NL_/day), or heavy (>30 g/day or >3 SU_NL_/day). Note that in our study, too few participants classified as very heavy drinkers (>60 g/day or >6 SU_NL_/day; n=129, 0.02%) to support definition of a sixth category. As a second indicator of habitual alcohol intake, we classified participants as binge-drinker if they reported drinking five or more drinks on a single occasion at least once in the past month (four or more for females), and non-binge-drinker if they did not [29]. Abstinent participants were included in the latter group. For descriptive purposes, we assessed each participant’s predominant drink type as the drink type that constituted >50% of their reported alcohol intake in grams per day. When no single drink type constituted >50%, drink type was classified as ‘mixed’.

### Outcome measure: executive functioning

Executive functioning was assessed with the Ruff Figural Fluency Test (RFFT). The RFFT is an overall measure of executive functioning comprising planning and reasoning, mental flexibility, inhibition, strategy generation and regulation of action [30]. The RFFT consists of five parts, each containing 35 five-dot patterns arranged in five columns and seven rows. Each part uses either different distractors or different patterns. Participants are instructed to draw as many unique designs between the dots as possible during sixty seconds. The total number of unique designs (range 0-175) was used as the dependent variable in the analyses, with higher scores representing better performance. Participants who failed to produce a single design throughout the test were excluded. The RFFT has a good test-retest and interrater reliability [31], [32], and is sensitive to age-related changes within the normal range [33]. Due to logistical reasons, the RFFT was administered in all Lifelines participants until April 2012, and in a random half of all participants thereafter.

### Potential confounders

We measured the following potential confounders: Demographic factors: age in years, sex, ethnicity (white vs. other); socioeconomic factors: neighbourhood status score by postcode (Dutch equivalent of the Townsend deprivation index [34]), monthly household income in euros, monthly household income equivalised, educational attainment (low, middle, high), occupational status (ISEI08 [35]); lifestyle factors: body mass index (BMI) in kg/m^2^, smoking (never, former, current), overall diet quality approximated by the Lifelines Diet Score [28], leisure-time and commuting moderate-to-vigorous physical activity in minutes per week, sleep duration relative to age and sex (short, normal, long); health factors: lifetime diagnosis of cardiovascular disease (yes/no), cancer (yes/no), diabetes (yes/no) or rheumatoid arthritis/liver cirrhosis (yes/no), current depressive disorder (yes/no) or anxiety disorder (yes/no), familial risk of cardiovascular disease (yes/no), depression/anxiety (yes/no) or addiction (yes/no); personality factors: NEO personality inventory facet sum score for impulsivity, excitement-seeking and self-discipline [36], past month estimated number of social contacts; and stress-related factors: past year number of stressful life events and long-term difficulties.

### Statistical procedures

All analyses were performed in R version 3.5.2. First, missing data points (degree of missingness ranging from <0.1% for BMI to 20.6% for diet quality) were imputed using multivariate imputation by chained equations [37], applying classification tree predictions for categorical variables and regression tree predictions for continuous variables. Categorical values were derived after imputation of the underlying continuous variables.

To estimate unadjusted associations between drinking level and all covariates, drinking level was treated as a pseudo-continuous variable ranging from 0 (abstinent) to 4 (>30 g/day). We estimated Pearson’s correlations between continuous variable pairs, polyserial correlations between continuous-categorical variable pairs, and polychoric correlations between categorical variable pairs. Similarly, unadjusted correlations were estimated between binge-drinking (yes/no) and each covariate. Significant (p<0.05) unadjusted associations ≥0.1 (absolute) are reported.

We applied a multinomial propensity score weighing approach to increase comparability of participants with different drinking levels, and of binge-drinking and non-binge-drinking participants. Propensity scores, designed to adjust incomplete randomization in intervention studies, have proven effective in removing selection bias in observational studies [38]. In two propensity models with average treatment effect (ATE) estimands, we predicted drinking level (abstinent/occasional/light/moderate/heavy) and binge-drinking (yes/no) from all covariates listed above and all possible two-way interactions between them. Multinomial propensity score estimation treats drinking levels as dummy variables, i.e. does not impose an order or linear dependence between levels. Contributions of individual confounders to each propensity score are found in Supplement 2. Residual imbalance between any two propensity-weighted groups not exceeding an absolute standardized mean difference (ASMD) of 0.1 was deemed acceptable, and ASMD of 0.1-0.25 was deemed adjustable [39].

In the analysis stage, propensity weighing is preferred over propensity score matching when comparing multiple unequally-sized groups, as it allows analysis of the full cohort and outperforms regression adjustment for propensity scores in removing bias [40]. We thus compared executive functioning between abstinent participants and participants of the other four drinking levels, and between binge-drinking and non-binge-drinking participants, applying the inverse probability of treatment weights. To address residual confounding while maintaining covariate balance, all covariates included in the propensity score models were included in the regression models as well (double-adjustment) [39].

Three sensitivity analyses were planned to assess robustness of our findings. First, propensity score estimation and weighted regression modelling were repeated in age- and sex-stratified samples, as both executive functioning and drinking patterns differ by age and sex [33], [41]. Age was stratified by tertiles (young: age 18-38; middle-age: 39-48; old: 48-65). Second, analyses were repeated drinkers of predominantly beer and wine separately, as drinking habits have been shown to differ by drink type even if total alcohol intake remains unchanged [42]. Drink types other than beer and wine were rarely predominant and could not be tested separately. Finally, analyses were repeated in participants of white ethnicity only, as ethnicity has been shown a particularly strong predictor of drinking habits in the Dutch population [43].

Standardised effect sizes (β) between 0.20 and 0.39 are generally referred to as ‘weak’. However, in our sample of 78,832 participants, with a power of 0.9 and α=0.010 (conservatively adjusted for testing one predictor with five levels and one binary predictor: 0.05/(5-1)+(2-1)=0.010), associations weaker than β<=0.05 (i.e. <5% change in the outcome variable with every standard deviation change in the predictor variable) may reach significance [44]. In well-powered models predicting complex behaviours, large βs are not expected and small βs can be informative [45]. Differentiation between statistically significant and clinically relevant findings is not achieved by further lowering of α, but rather by investigating β. Consensus regarding denotation of weak associations is lacking. Here, for interpretational purposes, we qualified absolute βs of statistically significant associations (p<0.01) between 0.05-0.19 as very weak, 0.2-0.39 as weak, 0.4-0.59 as moderate, 0.6-0.79 as strong and ≥0.8 as very strong. Associations with an absolute β<0.05 were considered to be non-informative even if they reached statistical significance.

## Results

### Descriptive

N=17,373 participants (22.0%) had been abstinent in the past month, compared to n=16,838 occasional drinkers (21.4%), n=33,827 light drinkers (42.9%), n=8,955 moderate drinkers (11.4%) and n=1,839 heavy drinkers (2.3%). A total of 8,380 participants (10.6%) had engaged in binge-drinking. Among drinkers, red wine was most often the predominant drink type (n=26,999 or 43.9%), followed by beer (n=10,993 or 17.9%), white wine (n=4,487 or 7.3%), and liquor (n=2,858 or 4.7%). The remaining drink types (alcohol-free beer, fortified wine, and other alcoholic drinks) were predominant in less than four percent of the sample. N=14,100 (22.9%) reported no predominant drink type (‘mixed’).

In unadjusted analyses, higher drinking levels were associated with male sex (r_FEMALE_=-0.393), excitement-seeking (r=0.210), current smoking (r_CURRENT-VS-NEVER_=0.144), higher equivalised income (r=0.133), white ethnicity (r_OTHER_=-0.125) and lower odds of depression (r=-0.113). Binge-drinking was also associated with male sex (r_FEMALE_=-0.239), current smoking (r_CURRENT-VS-NEVER_=0.295) and excitement seeking (r=0.334), but also with younger age (r=-0.307), lower diet quality (r=-0.277), impulsivity (r=0.183), lower odds of being a past smoker (r_PAST-VS-NEVER_=-0.125), lower income (r=-0. 149), lower educational attainment (r_HIGH-VS-MIDDLE_=-0.122), lower neighbourhood status (r=-0.113), lower occupational status (r=-0.111), and lower odds of cancer (r=-0.100) and cardiovascular disease (r=-0.112).

### Balance after weighing

Supplement 3, available online, provides the differences in sample characteristics before and after weighing was applied. Weighing reduced all ASMDs between binge-drinkers and non-binge-drinkers to acceptable levels, and all pairwise ASMDs between abstinent and other drinking levels to acceptable or adjustable levels (Supplemental Figure S3.4). Adjustable residual imbalance between groups involved almost exclusively the abstinent vs. heavy drinking contrast, and ranged from ASMD=0.11 (31% past-smokers in the abstinent group compared to 36% past-smokers in the heavy drinking group) to ASMD=0.24 (49% never-smokers in the abstinent group compared to 37% never-smokers in the heavy drinking group).

### Average Treatment Effects

After propensity score weighing, light drinkers (2.5-14.9 g/day) scored on average, 1.3 points higher on the executive functioning task compared to abstinent participants, indicating better performance (Table 1). Moderate drinkers (15-29.9 g/day) scored on average 2.5 points higher than abstinent participants. Executive functioning task performance did not differ between abstinent participants and occasional drinkers, or between abstinent participants and heavy drinkers. We found no difference in executive functioning task performance between binge-drinkers and non-binge-drinkers.

**Table 1.** Propensity weighted models comparing abstinent participants to each drinking level, and binge-drinkers to non-binge-drinkers

|  | Non-standardised |  | Standardised |  | p |
| --- | --- | --- | --- | --- | --- |
| | Estimate | 95% CI | $\beta$ | 95% CI | |
| Occasional (0-2.5 g/day) vs. abstinent | 0.415 | -0.141 – 0.971 | 0.018 | -0.006 – 0.043 | 0.1432 |
| Light (2.5-14.9 g/day) vs. abstinent | 1.264 | 0.755 – 1.773 | 0.056 | 0.033 – 0.078 | <0.0001 |
| Moderate (15-29.9 g/day) vs. abstinent | 2.521 | 1.792 – 3.249 | 0.111 | 0.079 – 0.143 | <0.0001 |
| Heavy (30-60 g/day) vs. abstinent | 1.689 | -0.196 – 3.575 | 0.075 | -0.009 – 0.158 | 0.0791 |
| Binge-drinking yes vs no | 0.722 | -0.046 – 1.490 | 0.032 | -0.002 – 0.066 | 0.0654 |

### Planned sensitivity analyses

Results were attenuated but remained significant when excluding participants of non-white ethnicity (Supplement 4). To assess robustness of our findings, propensity score estimation and weighted regression modelling were repeated in age- and sex-stratified samples. For a full description and results, see Supplement 5. As individual strata were insufficiently powered to allow detection of significant effects of the magnitude found in the full sample, we inspected effect sizes. The increased executive functioning task performance in light drinkers compared to abstinent participants, as seen in the full sample, was observed in all female strata (β ranging from 0.059 to 0.108) and in males of middle-age (β=0.093), but not in younger (β=-0.002) or older males (β=0.005). The increased task performance in moderate drinkers compared to abstinent participants was found in all male and female strata (β=0.052-0. 174) except in younger males (β=0.033). Note that the young males abstinent reference category suffered low N.

Propensity score estimation and weighted regression modelling were repeated comparing drinkers of predominantly beer (n=10,993) and drinkers of predominantly wine (n=39,139) to abstinent participants (Supplement 6). Better task performance among light drinking individuals, as found in the full sample, replicated in drinkers of wine (β[CI]=0.065[0.043-0.087]) but not in drinkers of beer (β[CI]=0.037[-0.010-0.085]). Better task performance among moderate drinkers replicated in both groups (beer: β[CI]=0.134[0.061-0.207]; wine: β[CI]=0.119[0.080-0.157]). However, propensity weighing could not achieve satisfactory balance between abstinent participants and light/moderate drinkers of beer, warranting cautious interpretation of findings regarding this group.

## Discussion

In an unprecedented sample of 78,832 generally healthy Dutch adults, we investigated associations between habitual levels of alcohol consumption and higher-order cognitive functioning. Applying propensity score weighing to adjust for a comprehensive set of demographic, socioeconomic, lifestyle, psychosocial and health-related factors influencing drinking behaviour, we found that light and moderate drinkers (consuming <3 drinks per day) scored higher on an executive functioning task compared to participants who consumed no alcohol. Both effects held for women of all ages, men age 39-48 and drinkers of predominantly wine, but not for men age <39. No significant effects were found comparing occasional drinkers or heavy drinkers to abstainers, or comparing binge-drinkers to non-binge-drinkers.

The observed association between alcohol consumption and executive functioning was dose-dependent in a non-linear fashion, with no effect for occasional drinking (<2.5 g/day), significant positive effects for light (2.5-14.9 g/day) and moderate drinking (15-29.9 g/day), and a non-significant positive effect for heavy drinking (>30 g/day). Importantly, and contrary to prior studies, all our analyses were conditioned on a broad range of socioeconomic confounders (i.e. income, household size, educational attainment, neighbourhood deprivation) and lifestyle factors (i.e. diet quality, smoking, physical activity, sleep) that are associated with drinking. Two considerations are warranted. First, standardised effect sizes in our study never exceeded β=0.2, indicating that any observed effect, although significant, was very weak in strength. Second, the non-significant estimate in our heavy drinking group may reflect the smaller number of participants in this group (2.3%). Our findings thus support the left side of the inverted J-shape, but provides only preliminary evidence to support its right side.

The inverted J-shaped pattern resembles that of the relative risk of incident coronary heart disease at different drinking levels compared to abstinence [5], tentatively suggesting that cardiovascular factors might mediate the association between moderate alcohol consumption and better executive functioning. Cardiovascular effects of moderate alcohol consumption include protective effects such as lower blood pressure [7] and elevated high-density lipoprotein cholesterol [46] and detrimental effects such as glucose- and metabolic dysregulation [47]. Among these, lower blood pressure has most consistently been associated with better cognitive performance [48], [49], and may thus be a likely mediator. In addition, moderate alcohol consumption induces a reduction of inflammatory marker C-reactive protein (CRP) [50], a non-traditional risk factor for cardiovascular disease [51]. Lower CRP, in turn, has also been associated with better executive functioning [52], [53]. Alternative study designs are needed to elucidate the potential mediating role of cardiovascular and inflammatory factors, as our cross-sectional data does not support testing of mediation hypotheses. Nonetheless, introducing blood pressure and/or CRP as a covariate to our model did not alter the estimated association between alcohol consumption and executive functioning (data not shown), tentatively suggesting that other factors might be at play.

Intuitively, subtle brain changes resulting from alcohol consumption may be proposed as an alternative intermediate factor. Population-based neuroimaging studies, however, have convincingly shown negative associations between total brain volume and moderate alcohol consumption [54]–[57], suggesting detrimental rather than beneficial effects of even moderate drinking. In fact, the one study performing regional analyses reported that lifetime alcohol intake was associated with smaller volume of the middle frontal cortex [56], an area crucially implicated in executive control [58]. The observation of improved cognitive outcomes in the presence of unfavourable brain changes has been described as the “alcohol paradox” [59]. We conclude that the association between moderate drinking and better executive functioning is unlikely to be mediated by volumetric brain changes. However, other brain parameters (e.g. task-induced activity, structural or functional connectivity) may also be of interest.

We wish to bring to the reader’s attention two purely methodological explanations of our finding of better executive functioning among light/moderate drinkers. First, it has frequently been proposed that individuals who refrain from drinking for health reasons, including former drinkers or ‘sick quitters’, may drive poorer outcomes in the abstinent group. Former drinking was not specifically assessed in our sample. However, our propensity metrics included somatic, mental and familial health factors that may persuade individuals to refrain from drinking. In more general terms, the influence of sick quitters is often overestimated, as they make up a relatively small proportion of all abstainers and removing former drinkers from the analyses typically has minimal impact on group statistics [60]. Second, one might argue that the positive association between drinking and executive functioning may suggest collider bias. Collider bias occurs when conditioning upon a third variable, that is influenced by both the predictor and the outcome, causes a spurious association between the predictor and the outcome. We exclude the possibility of a measured collider variable, as unadjusted associations similarly produced positive associations between light/moderate drinking and task performance (data not shown). Unmeasured colliders can occur through selection bias [61]. Note, however, that our significant contrasts involved light/moderate drinkers rather than heavy/problematic drinkers (the latter group being more susceptible to selection bias for health reasons), and that the Lifelines sample is generally representative of the northern Netherlands population [62]. Thus, although the sick-quitter- and collider-hypotheses cannot fully be disproved, we conclude that both explanations are rather unlikely.

Three limitations of our work should be kept in mind. First, in the above interpretations of our cross-sectional findings, we assumed that drinking level affects executive functioning task performance rather than the other way around. Balancing drinking groups based on factors predicting alcohol intake (instead of executive functioning) reinforces this assumption, but does not fully eliminate the possibility that reverse causation might occur. Specifically, we cannot exclude the possibility that light/moderate drinkers may not progress into heavy/problematic drinking due to their effective higher-order cognitive skills. Second, executive functioning was assessed using a single figural fluency task, which correlates with – but does not fully capture – other executive domains such as inhibitory control, cognitive flexibility and working memory [31], [63]. Drinking may be differentially associated with other executive domains. Finally, third, due to the observational nature of our study, we cannot exclude the possibility of residual (unmeasured, potentially genetic) confounding. The partial attenuation of effects after excluding participants of non-white ethnicity suggests that residual confounding might contribute.

In sum, we observed better executive functioning task performance in light and moderate drinkers of alcohol compared to abstainers, but not in occasional drinkers, heavy drinkers or binge-drinkers. This non-linear association is reminiscent of the dose-dependent effect of alcohol consumption on cardiovascular risks. In a representative population-based sample ten times the size of the largest study to date, we addressed non-random group allocation and risk of confounding. Our work contributes to the notion that a ‘safe level’ of alcohol consumption may exist for some health outcomes but not for others (e.g. cancer, injury from falls), making formulation of universal guidelines extremely challenging. In the future, identifying (partial) mediators of the association between moderate drinking and higher-order cognitive functions may advance our understanding of underlying biological mechanisms.

## Data Availability

The data analyzed in this study was obtained from the Lifelines biobank (project application number OV15_0307). Requests to access this dataset should be directed to Lifelines Research Office.

https://www.lifelines.nl/

## Conflicts of interest

During the past three years, JH has received educational speaking fees from HB Pharma, Biocodex, Medice, Takeda and Shire. H Larsson has served as a speaker for Evolan Pharma and Shire/Takeda and has received research grants from Shire/Takeda; all outside the submitted work. The other authors declare no conflicts of interest.

## Notes

### Funding Statement

This work was supported by the European Unions Horizon 2020 Research and Innovation Program under grant agreement No 728018. The funding source has had no involvement in the study design, data collection, interpretation of the findings, or writing of this manuscript.

### Author Declarations

The Lifelines Cohort Study is conducted according to the principles of the Declaration of Helsinki and is approved by the medical ethics committee of the University Medical Center Groningen, The Netherlands.

