## Supplemental Materials for "Drinking habits and executive functioning: a propensity score-weighted analysis of 78,832 adults"

Contents:


1. Participant flowchart
2. Heatmap: propensity scores vs. all potential confounders
3. Balance before and after propensity score weighing
    i. Means by drinking level before and after weighing
    ii. Means by binge-drinking (yes/no) before and after weighing
    iii. Adjustable and non-adjustable differences between weighted groups
4. Results among participants of white ethnicity only
5. Results stratified by age and sex
6. Results stratified by drink type


PRE-PRINT

### Supplement 1: Participant flowchart

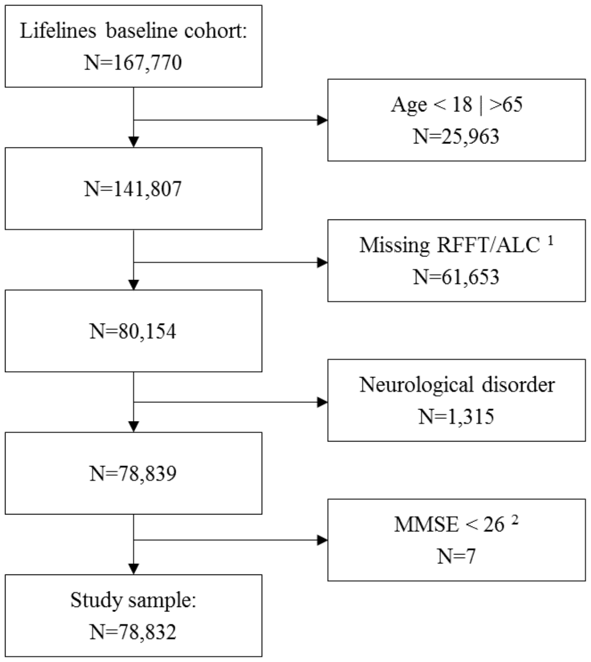


**Supplemental Figure S1**. Participant flowchart.

RFFT = Ruff Figural Fluency Task; MMSE = Mini Mental State Examination; 1 For logistical reasons, the RFFT was administered in all Lifelines participants until April 2012, and in a random half of all participants thereafter, resulting in a relatively large proportion of missing data; 2 By protocol, the RFFT was not administered in participants with MMSE<26. Moreover, MMSE was only administered in participants age 65 and older. Thus, the vast majority of participants with MMSE<26 were already excluded in a previous step.

PRE-PRINT

### Supplement 2: Correlations between propensity scores and all potential confounders

Correlation (-1.0, -0.5, 0.0, 0.5, 1.0)

| Domain | Confounder | abstinent | occasional | light | moderate | heavy | binge |
|---|---|---|---|---|---|---|---|
| ***Demographic*** | age | 0.02 | -0.29 | -0.07 | 0.26 | 0.08 | -0.49 |
| | sex | 0.66 | 0.67 | -0.61 | -0.7 | -0.57 | -0.39 |
| | ethnicity | 0.21 | 0.26 | -0.26 | -0.15 | -0.12 | -0.04 |
| ***SES*** | low educational level | 0.17 | -0.12 | -0.21 | 0.04 | 0.1 | 0.07 |
| | high educational level | -0.2 | 0.03 | 0.25 | 0.05 | -0.08 | -0.13 |
| | income | -0.2 | -0.12 | 0.24 | 0.16 | -0.01 | -0.24 |
| | income (eq) | -0.27 | -0.18 | 0.28 | 0.24 | 0.05 | -0.06 |
| | neighbourhood status | 0 | -0.04 | 0.04 | 0.01 | -0.06 | -0.18 |
| | occupational status | -0.22 | -0.02 | 0.27 | 0.1 | -0.07 | -0.17 |
| ***Health*** | BMI | 0.19 | -0.01 | -0.26 | -0.03 | 0.1 | 0.05 |
| | cancer | 0.01 | 0.04 | -0.05 | 0.01 | -0.01 | -0.15 |
| | cardiovascular | 0.13 | -0.15 | -0.17 | 0.07 | 0.1 | -0.16 |
| | diabetes | 0.26 | -0.05 | -0.31 | -0.03 | 0.01 | -0.1 |
| | liver cirrhosis | 0.14 | -0.06 | -0.17 | 0.01 | 0.04 | -0.09 |
| | depression | 0.29 | 0.02 | -0.31 | -0.12 | 0.02 | 0.06 |
| | anxiety | 0.2 | 0.05 | -0.26 | -0.07 | 0.01 | 0 |
| ***Family health*** | addiction | 0.02 | -0.12 | -0.03 | 0.06 | 0.08 | 0 |
| | depression/anxiety | 0.03 | 0.05 | -0.02 | -0.04 | -0.04 | -0.12 |
| | CVD | 0.09 | -0.03 | -0.09 | -0.01 | -0.01 | -0.07 |
| ***Lifestyle*** | diet quality | 0 | 0.13 | 0.15 | -0.17 | -0.33 | -0.43 |
| | physical activity | -0.07 | -0.06 | 0.12 | 0.02 | -0.02 | 0.02 |
| | past smoking | -0.15 | -0.22 | 0.18 | 0.21 | 0.02 | -0.14 |
| | current smoking | -0.11 | -0.32 | 0.01 | 0.33 | 0.3 | 0.38 |
| | short sleep | 0.04 | -0.03 | -0.09 | 0.03 | 0.06 | 0.01 |
| | long sleep | 0.08 | 0.07 | -0.12 | -0.04 | 0.01 | -0.01 |
| ***Stress*** | stressful life events | 0.07 | 0 | -0.12 | 0 | 0.07 | 0.08 |
| | long-term stress | 0.05 | 0.16 | -0.1 | -0.08 | 0 | 0.08 |
| ***Personality*** | impulsivity | -0.07 | -0.04 | 0.04 | 0.07 | 0.11 | 0.29 |
| | excitement-seeking | -0.43 | -0.21 | 0.47 | 0.26 | 0.19 | 0.52 |
| | self-discipline | -0.03 | -0.09 | 0.09 | 0.03 | -0.03 | -0.11 |
| | social contacts | -0.07 | -0.17 | 0.13 | 0.06 | 0.05 | 0.05 |

**Supplemental Figure S2.** Heatmap of correlations between all confounders and each propensity score.

### Supplement 3: Balance before and after weighing

**Supplemental Table S3.1.** Means by drinking level before and after weighing

| | Before weighing | | | | | After weighing | | | | |
|---|---|---|---|---|---|---|---|---|---|---|
| | abst. | occ. | light | mod. | heavy | abst. | occ. | light | mod. | heavy |
| Age | 42.9 | 41.7 | 42.6 | 45.1 | 44.4 | 43.2 | 42.8 | 42.9 | 43.5 | 44.4* |
| Income | 2309.8 | 2407.9 | 2498.1 | 2543.6 | 2415.4 | 2432.6 | 2442.1 | 2448.2 | 2469.6 | 2505.2 |
| Income equivalised | 1483.3 | 1561.1 | 1636.1 | 1694.3 | 1643.2 | 1586.7 | 1589.1 | 1598.5 | 1618.0 | 1665.4* |
| Neighbourhood status | -0.6 | -0.6 | -0.6 | -0.6 | -0.7 | -0.6 | -0.6 | -0.6 | -0.6 | -0.6 |
| Occupational status | 42.4 | 46.0 | 47.8 | 47.8 | 43.5 | 46.0 | 46.1 | 46.3 | 46.7 | 46.4 |
| BMI | 26.7 | 26.0 | 25.7 | 25.9 | 26.8 | 26.1 | 26.0 | 26.0 | 25.9 | 26.1 |
| Diet quality | 23.7 | 23.9 | 23.9 | 23.0 | 20.4 | 23.7 | 23.7 | 23.7 | 23.6 | 23.1* |
| Physical activity | 249.3 | 259.1 | 275.0 | 270.2 | 254.9 | 259.1 | 260.9 | 264.3 | 257.4 | 259.8 |
| SLE | 1.3 | 1.2 | 1.1 | 1.2 | 1.4 | 1.2 | 1.2 | 1.2 | 1.2 | 1.2 |
| LTD | 2.7 | 2.7 | 2.5 | 2.4 | 2.6 | 2.6 | 2.6 | 2.6 | 2.6 | 2.5 |
| Impulsivity | 22.1 | 22.3 | 22.4 | 22.6 | 23.2 | 22.3 | 22.3 | 22.4 | 22.4 | 22.6 |
| Excitement seeking | 20.5 | 21.8 | 22.9 | 23.2 | 23.9 | 22.0 | 22.1 | 22.3 | 22.3 | 22.6* |
| Self-discipline | 29.2 | 29.1 | 29.4 | 29.4 | 29.0 | 29.3 | 29.3 | 29.3 | 29.3 | 29.5 |
| Social contacts | 16.7 | 16.7 | 18.1 | 18.2 | 18.7 | 17.3 | 17.4 | 17.5 | 17.3 | 17.9 |
| Male sex(%) | 19.4 | 31.8 | 48.3 | 63.5 | 82.2 | 41.6 | 40.0 | 42.1 | 45.4 | 51.3* |
| Nonwhite ethnicity (%) | 2.1 | 1.8 | 1.1 | 0.9 | 0.5 | 1.6 | 1.5 | 1.4 | 1.1 | 1.1 |
| Low education (%) | 34.9 | 26.6 | 25.6 | 29.7 | 37.5 | 29.3 | 28.6 | 28.4 | 28.8 | 29.6 |
| High education (%) | 22.7 | 30.7 | 22.8 | 32.5 | 23.2 | 28.9 | 30.2 | 30.6 | 31.2 | 30.2 |
| Cancer (%) | 3.7 | 3.8 | 3.6 | 3.7 | 3.5 | 3.7 | 3.8 | 3.8 | 3.5 | 4.7 |
| CVD (%) | 34.3 | 28.5 | 28.6 | 32.4 | 37.1 | 31.2 | 30.3 | 30.3 | 30.6 | 33.3 |
| Diabetes (%) | 2.9 | 1.8 | 1.4 | 1.7 | 1.8 | 2.1 | 1.9 | 1.8 | 2.2 | 1.6 |
| Liver cirrhosis (%) | 2.5 | 1.8 | 1.7 | 1.9 | 2.2 | 2.0 | 1.9 | 1.8 | 1.9 | 1.3 |
| Depression (%) | 5.0 | 3.2 | 2.5 | 2.5 | 3.3 | 3.3 | 3.1 | 3.1 | 3.1 | 2.9 |
| Anxiety (%) | 10.7 | 8.3 | 6.9 | 7.3 | 8.1 | 8.2 | 8.1 | 7.9 | 8.7 | 7.7 |
| Fam addiction (%) | 5.6 | 4.9 | 5.3 | 6.0 | 7.3 | 5.8 | 5.3 | 5.4 | 5.7 | 5.6 |
| Fam dep/anx (%) | 27.0 | 26.8 | 26.1 | 25.0 | 24.1 | 26.2 | 26.4 | 26.1 | 25.7 | 26.7 |
| Fam CVD (%) | 10.7 | 9.4 | 8.9 | 9.2 | 8.9 | 9.9 | 9.5 | 9.4 | 8.9 | 9.7 |

PREPRINT

| | | | | | | | | | | |
|---|---|---|---|---|---|---|---|---|---|---|
| Past smoking (%) | 25.8 | 28.1 | 34.3 | 40.0 | 34.5 | 30.9 | 31.8 | 32.1 | 33.8 | 36.0* |
| Current smoking (%) | 17.7 | 17.1 | 22.1 | 33.3 | 44.9 | 20.4 | 21.3 | 22.4 | 23.4 | 27.2* |
| Short sleep (%) | 11.0 | 9.7 | 9.2 | 10.9 | 13.7 | 10.3 | 10.1 | 10.1 | 10.7 | 10.0 |
| Long sleep (%) | 12.0 | 10.8 | 8.8 | 8.8 | 10.4 | 10.2 | 10.1 | 9.7 | 9.3 | 9.6 |

* adjustable difference between indicated group and abstinent group after weighing. Abbreviations: abst. = abstinent, occ. = occasional drinker, mod. = moderate drinker.

**Supplemental Table S3.2.** Means by binge-drinking (yes/no) before and after weighing

| | Before weighing | | After weighing | |
|---|---|---|---|---|
| | non-binge-drinking | binge-drinking | non-binge-drinking | binge-drinking |
| Age | 43.5 | 36.8 | 42.8 | 42.6 |
| Income | 2466.3 | 2224.1 | 2442.9 | 2444.1 |
| Income equivalised | 1597.5 | 1557.0 | 1593.6 | 1608.2 |
| Neighbourhood status | -0.6 | -0.8 | -0.6 | -0.6 |
| Occupational status | 46.6 | 42.2 | 46.2 | 45.5 |
| BMI | 26.0 | 26.2 | 26.0 | 26.1 |
| Diet quality | 24.0 | 21.0 | 23.7 | 23.4 |
| Physical activity | 264.2 | 270.6 | 264.6 | 267.4 |
| SLE | 1.2 | 1.3 | 1.2 | 1.2 |
| LTD | 2.6 | 2.8 | 2.6 | 2.6 |
| Impulsivity | 22.2 | 23.6 | 22.4 | 22.5 |
| Excitement seeking | 21.9 | 24.9 | 22.2 | 22.5 |
| Self-discipline | 29.3 | 28.7 | 29.3 | 29.4 |
| Social contacts | 17.4 | 18.3 | 17.5 | 17.2 |
| Male sex(%) | 39.0 | 57.4 | 40.8 | 43.3 |
| Nonwhite ethnicity (%) | 1.5 | 1.2 | 1.5 | 1.4 |
| Low education (%) | 28.2 | 32.4 | 28.6 | 29.3 |
| High education (%) | 31.1 | 23.2 | 30.3 | 28.1 |
| Cancer (%) | 3.8 | 2.5 | 3.7 | 3.8 |
| CVD (%) | 31.2 | 23.9 | 30.5 | 30.3 |
| Diabetes (%) | 1.9 | 1.3 | 1.9 | 1.7 |
| Liver cirrhosis (%) | 2.0 | 1.4 | 1.9 | 1.6 |
| Depression (%) | 3.1 | 3.7 | 3.2 | 3.1 |

PRE-PRINT

| | | | | |
|---|---|---|---|---|
| Anxiety (%) | 8.1 | 7.9 | 8.1 | 8.3 |
| Fam addiction (%) | 5.4 | 5.5 | 5.4 | 5.7 |
| Fam dep/anx (%) | 26.8 | 21.8 | 26.3 | 26.0 |
| Fam CVD (%) | 9.6 | 8.0 | 9.5 | 9.0 |
| Past smoking (%) | 32.6 | 24.4 | 31.8 | 33.9 |
| Current smoking (%) | 19.8 | 39.2 | 21.8 | 23.9 |
| Short sleep (%) | 10.0 | 10.3 | 10.0 | 9.6 |
| Long sleep (%) | 10.0 | 9.5 | 10.0 | 9.9 |

**Supplemental Table S3.3.** Adjustable and non-adjustable differences between weighted groups

| | Contrast | Confounder | Group 1 | Group 2 | ASMD |
|---|---|---|---|---|---|
| Adjustable | Abstinent vs. moderate | Smoking: never | 48.7% | 42,8% | 0.1185 |
| | Abstinent vs. heavy | Age | 43.2 | 44.4 | 0.1103 |
| | Abstinent vs. heavy | Sex: male | 41.6% | 51.3% | 0.1946 |
| | Abstinent vs. heavy | Income equivalised | 1586.7 | 1665.4 | 0.1553 |
| | Abstinent vs. heavy | Diet quality | 23.7 | 23.1 | 0.1148 |
| | Abstinent vs. heavy | Smoking: never | 48.7% | 36.8% | 0.2385 |
| | Abstinent vs. heavy | Smoking: past | 30.9% | 36.0% | 0.1066 |
| | Abstinent vs. heavy | Smoking: current | 20.4% | 27.2% | 0.1614 |
| | Abstinent vs. heavy | Excitement-seeking | 22.0 | 22.6 | 0.1404 |
| Non-adjustable | *None* | | | | |

PREPRINT

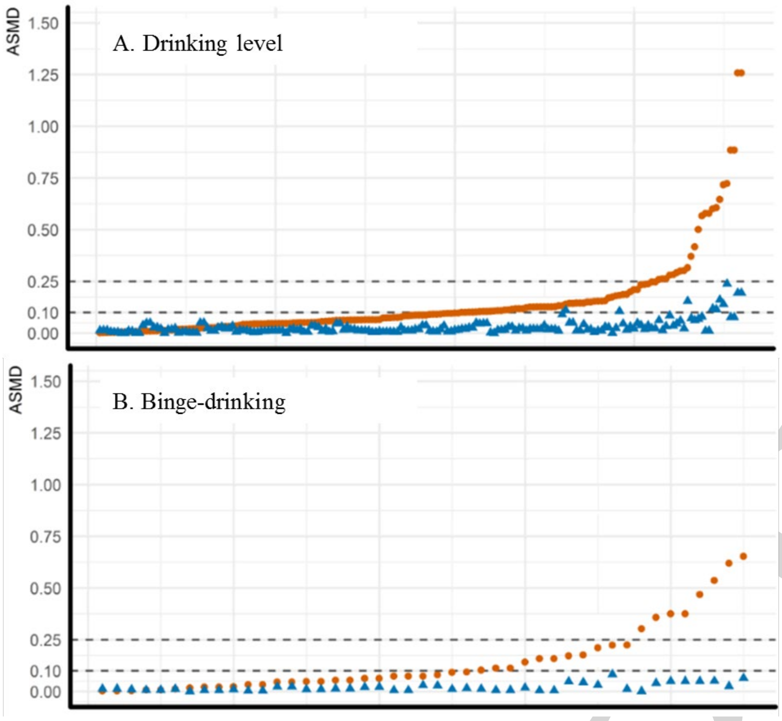


**Supplemental Figure S3.4**. Absolute standardised mean differences (ASMD) in pairwise between-group contrasts (e.g. $ASMD_{AGE}$ abstinent vs. light drinkers, $ASMD_{AGE}$ abstinent vs. moderate drinkers, etc.) before (red circle) and after (blue triangle) propensity score weighing. On the X-axis, contrasts are ordered by their pre-weighing ASMD. Dashed horizontal lines indicate acceptable (ASMD<0.1) and adjustable (ASMD<0.25) residual imbalance after weighing. No residual between-group difference exceeds ASMD=0.25.

PRE-PRINT

### Supplement 4: Results among participants of white ethnicity only

**Supplemental Table S4.1**. Adjustable and non-adjustable differences between weighted groups

| | Contrast | Confounder | Group 1 | Group 2 | ASMD |
|---|---|---|---|---|---|
| Adjustable | Abstinent vs. moderate | Smoking: never | 48.4% | 43.3% | 0.1016 |
| | Abstinent vs. heavy | Diet quality | 23.7 | 23.0 | 0.1320 |
| | Abstinent vs. heavy | Smoking: never | 48.4% | 40.9% | 0.1502 |
| | Abstinent vs. heavy | Smoking: current | 20.6% | 25.5% | 0.1165 |
| | Abstinent vs. heavy | Excitement-seeking | 21.9 | 22.7 | 0.1549 |
| Non-adjustable | *None* | | | | |

**Supplemental Table S4.2**. Propensity weighted models comparing abstinent participants to each drinking level, and binge-drinkers to non-binge-drinkers

| | Non-standardised | | Standardised | | |
|---|---|---|---|---|---|
| | Est | 95% CI | β | 95% CI | p |
| Occasional (0-2.5 g/day) vs. abstinent | 0.343 | -0.179 – 0.864 | 0.015 | -0.008 – 0.038 | 0.1974 |
| Light (2.5-14.9 g/day) vs. abstinent | 0.849 | 0.389 – 1.318 | 0.038 | 0.017 – 0.058 | 0.0004 |
| Moderate15-29.9 g/day) vs. abstinent | 1.793 | 1.122 – 2.465 | 0.079 | 0.050 – 0.109 | <0.0001 |
| Heavy (30-60 g/day) vs. abstinent | 0.789 | -0.977 – 2.736 | 0.039 | -0.043 – 0.121 | 0.3531 |
| Binge-drinking yes vs no | 0.233 | -0.528 – 0.994 | 0.010 | -0.023 – 0.044 | 0.5479 |

PRE-PRINT

### Supplement 5: Results stratified by age and sex

**Supplemental Table S5.1**. Age- and sex-stratified groups

| Sex | Age | N | % |
|---|---|---|---|
| Male | Young (18-38) | 10,392 | 13.2 |
| | Middle (39-48) | 11,999 | 15.2 |
| | Old (49-65) | 9,876 | 12.5 |
| Female | Young (18-38) | 15,442 | 19.6 |
| | Middle (39-48) | 17,287 | 21.9 |
| | Old (49-65) | 13,836 | 17.6 |

**Supplemental Table S5.2**. Adjustable and non-adjustable differences between weighted groups

| | Group | Contrast | Confounder | Group 1 | Group 2 | ASMD |
|---|---|---|---|---|---|---|
| Adjustable | Male – young | Abst. vs. light | Excitement-seeking | 25.1 | 25.6 | 0.1217 |
| | | Abst. vs. moderate | Smoking: never | 55.3% | 48.4% | 0.1377 |
| | | Abst. vs. moderate | Excitement-seeking | 25.1 | 25.7 | 0.1582 |
| | | Abst. vs. heavy | Ethnicity: white | 98.4% | 99.7% | 0.1267 |
| | Male – middle | Abst. vs. heavy | Ethnicity: white | 98.7% | 99.9% | 0.1114 |
| | | Abst. vs. heavy | Excitement-seeking | 23.2 | 23.7 | 0.1172 |
| | Male – old | Abst. vs. moderate | Excitement-seeking | 21.5 | 21.9 | 0.1003 |
| | | Abst. vs. heavy | Ethnicity: white | 98.6% | 99.8% | 0.1252 |
| | | Abst. vs. heavy | Family: addiction | 5.3% | 3.1% | 0.1029 |
| | | Abst. vs. heavy | Excitement-seeking | 21.5 | 22.2 | 0.1688 |
| | Female – young | Abst. vs. light | Excitement-seeking | 22.9 | 23.3 | 0.1019 |
| | | Abst. vs. moderate | Education: middle | 49.1% | 43.7% | 0.1085 |
| | | Abst. vs. moderate | Education: high | 36.1% | 43.7% | 0.1532 |
| | | Abst. vs. moderate | BMI | 25.1 | 24.5 | 0.1281 |
| | | Abst. vs. moderate | Family: cardiovascular | 7.3% | 2.9% | 0.1832 |
| | | Abst. vs. moderate | Smoking: never | 60.2% | 52.3% | 0.1575 |
| | | Abst. vs. moderate | Smoking: current | 21.1% | 26.2% | 0.1164 |
| | | Abst. vs. moderate | Excitement-seeking | 22.9 | 23.7 | 0.1782 |

PRE-PRINT

| | | | | | | |
|---|---|---|---|---|---|---|
| | | Abst. vs. heavy | Sleep: short | 7.3% | 2.4% | 0.1904 |
| | | Abst. vs. heavy | Sleep: long | 12.7% | 5.2% | 0.2331 |
| | | Binge vs. non-binge | Age | 29.6 | 30.2 | 0.1032 |
| | | Binge vs. non-binge | Smoking: never | 52.2% | 57.2% | 0.1017 |
| | | Binge vs. non-binge | Excitement-seeking | 23.7 | 23.2 | 0.1302 |
| | Female – middle | Abst. vs. moderate | Income | 2500.8 | 2578.1 | 0.1008 |
| | | Abst. vs. moderate | Income (equivalised) | 1490.7 | 1552.2 | 0.1325 |
| | | Abst. vs. moderate | Smoking: never | 50.7% | 44.9% | 0.1166 |
| | | Abst. vs. moderate | Sleep: long | 9.9% | 6.6% | 0.1156 |
| | | Abst. vs. heavy | Ethnicity: white | 98.5% | 100.0% | 0.1276 |
| | | Binge vs. non-binge | Impulsivity | 22.9 | 22.5 | 0.1002 |
| | Female – old | Abst. vs. moderate | Income (equivalised) | 1651.2 | 1705.4 | 0.1055 |
| | | Abst. vs. moderate | Smoking: never | 37.2% | 31.8% | 0.1163 |
| | | Abst. vs. heavy | Ethnicity: white | 99.0% | 100.0% | 0.1076 |
| | | Abst. vs. heavy | Income | 2372.1 | 2525.2 | 0.1990 |
| | | Abst. vs. heavy | Income (equivalised) | 1651.2 | 1772.0 | 0.2350 |
| | | Abst. vs. heavy | Diabetes | 3.8% | 0.3% | 0.2096 |
| | | Abst. vs. heavy | Smoking: past | 46.4% | 57.8% | 0.2281 |
| | | Abst. vs. heavy | Smoking: current | 16.4% | 23.1% | 0.1787 |
| | | Binge vs. non-binge | Excitement-seeking | 20.0 | 19.5 | 0.1426 |
| Non-adjustable | Female – young | Abst. vs. heavy | Income | 2254.0 | 1777.0 | 0.5428 |
| | | Abst. vs. heavy | Income (equivalised) | 1514.8 | 1372.2 | 0.2681 |
| | | Abst. vs. heavy | Family: dep/anx | 28.5% | 10.0% | 0.4118 |
| | | Abst. vs. heavy | Sleep: normal | 80.0% | 92.4% | 0.3169 |
| | Female – middle | Abst. vs. heavy | Income (equivalised) | 1490.7 | 1609.7 | 0.2568 |
| | | Abst. vs. heavy | Cancer | 3.8% | 12.1% | 0.4224 |
| | Female – old | Abst. vs. heavy | Smoking: never | 37.2% | 19.1% | 0.3875 |

Abbreviations: abst. = abstinent

PREPRINT

**Supplemental Table S5.3**. Fully adjusted models comparing abstinent participants to each drinking level, and binge-drinkers to non-binge-drinkers, in age- and sex stratified groups

| | | N | β | 95% CI | p |
|---|---|---|---|---|---|
| Occasional (0-2.5 g/day) vs. abstinent | Full sample | 16,838 | 0.018 | -0.006 – 0.043 | 0.1432 |
| | Male – young | 1,710 | -0.018 | -0.109 – 0.074 | 0.7084 |
| | Male – middle | 2,181 | 0.044 | -0.031 – 0.119 | 0.2494 |
| | Male – old | 1,456 | -0.026 | -0.109 – 0.056 | 0.5329 |
| | Female – young | 4,487 | 0.024 | -0.022 – 0.069 | 0.3053 |
| | Female – middle | 4,142 | 0.045 | 0.004 – 0.086 | 0.0312 |
| | Female – old | 2,862 | 0.069 | 0.020 – 0.117 | 0.0056 |
| Light (2.5-14.9 g/day) vs. abstinent | Full sample | 33,827 | 0.056 | 0.033 – 0.078 | <0.0001 |
| | Male – young | 5,605 | -0.002 | -0.085 – 0.080 | 0.9541 |
| | Male – middle | 5,916 | 0.093 | 00.26 – 0.160 | 0.0065 |
| | Male – old | 4,821 | 0.005 | -0.067 – 0.077 | 0.8912 |
| | Female – young | 5,862 | 0.059 | 0.014 – 0.104 | 0.0104 |
| | Female – middle | 6,095 | 0.108 | 0.069 – 0.147 | <0.0001 |
| | Female – old | 5,528 | 0.106 | 0.064 – 0.149 | <0.0001 |
| Moderate (15-29.9 g/day) vs. abstinent | Full sample | 8,955 | 0.111 | 0.079 – 0.143 | <0.0001 |
| | Male – young | 1,695 | 0.033 | -0.061 – 0.127 | 0.4893 |
| | Male – middle | 1,966 | 0.174 | 0.098 – 0.250 | <0.0001 |
| | Male – old | 2,030 | 0.052 | -0.026 – 0.131 | 0.1917 |
| | Female – young | 584 | 0.131 | 0.024 – 0.238 | 0.0165 |
| | Female – middle | 1,094 | 0.145 | 0.067 – 0.224 | 0.0003 |
| | Female – old | 1,586 | 0.153 | 0.089 – 0.218 | <0.0001 |
| Heavy (30-60 g/day) vs. abstinent | Full sample | 1,839 | 0.075 | -0.009 – 0.158 | 0.0791 |
| | Male – young | 452 | 0.033 | -0.104 – 0.169 | 0.6389 |
| | Male – middle | 529 | 0.111 | -0.028 – 0.249 | 0.1168 |
| | Male – old | 531 | 0.054 | -0.063 – 0.171 | 0.3655 |
| | Female – young * | 40 | -0.401 | -0.740 – -0.062 | 0.0206 |
| | Female – middle * | 122 | 0.021 | -0.267 – 0.309 | 0.8869 |
| | Female – old * | 165 | 0.094 | -0.065 – 0.252 | 0.2484 |
| Binge-drinking yes vs no | Full sample | 8,380 | 0.032 | -0.002 – 0.066 | 0.0654 |
| | Male – young | 2,426 | 0.022 | -0.029 – 0.073 | 0.3940 |

PRE-PRINT

| | | | | | |
|---|---|---|---|---|---|
| | Male – middle | 1,534 | 0.021 | -0.038 – 0.080 | 0.4816 |
| | Male – old | 851 | -0.071 | -0.162 – 0.020 | 0.1249 |
| | Female – young | 2,111 | 0.036 | -0.028 – 0.101 | 0.2713 |
| | Female – middle | 909 | 0.134 | 0.046 – 0.222 | 0.0030 |
| | Female – old | 549 | 0.052 | -0.061 – 0.165 | 0.3670 |

PRE-PRINT

### Supplement 6: Results stratified by drink type

**Supplemental Table S6.1**. Adjustable and non-adjustable differences between weighted groups

| | Group | Contrast | Confounder | Group 1 | Group 2 | ASMD |
|---|---|---|---|---|---|---|
| Adjustable | Beer | Abst vs. occasional | Sex: male | 41.8% | 48.0% | 0.1698 |
| | | Abst vs. occasional | Smoking: never | 52.1% | 46.2% | 0.1202 |
| | | Abst vs. occasional | Smoking: current | 21.6% | 26.5% | 0.1068 |
| | | Abst vs. light | Smoking: never | 52.1% | 44.6% | 0.1523 |
| | | Abst vs. light | Smoking: current | 21.6% | 27.1% | 0.1196 |
| | | Abst vs. light | Excitement-seeking | 21.7 | 22.6 | 0.1884 |
| | | Abst vs. moderate | Diet quality | 22.8 | 22.0 | 0.1698 |
| | | Abst vs. moderate | Excitement-seeking | 21.7 | 22.8 | 0.2304 |
| | | Abst vs. heavy | Ethnicity: white | 98.2% | 100.0% | 0.1755 |
| | | Abst vs. heavy | Rheumatoid/liver disease | 2.2% | 0.7% | 0.1258 |
| | | Abst vs. heavy | Diet quality | 22.8 | 21.8 | 0.2017 |
| | | Binge vs. non-binge | Age | 40.6 | 41.8 | 0.1159 |
| | | Binge vs. non-binge | Sex: male | 55.1% | 44.4% | 0.2149 |
| | | Binge vs. non-binge | Diet quality | 22.2% | 22.8% | 0.1111 |
| | | Binge vs. non-binge | Smoking: never | 43.1% | 51.1% | 0.1585 |
| | | Binge vs. non-binge | Excitement-seeking | 22.7 | 21.9 | 0.1758 |
| | Wine | Abst vs. moderate | Smoking: never | 49.4% | 43.8% | 0.1122 |
| | | Abst vs. heavy | Age | 43.6 | 45.9 | 0.2185 |
| | | Abst vs. heavy | Sex: male | 30.1% | 38.6% | 0.1819 |
| | | Abst vs. heavy | Smoking: past | 32.1% | 39.8% | 0.1607 |
| | | Abst vs. heavy | Smoking: current | 18.5% | 25.7% | 0.1795 |
| Non-adjustable | Beer | Abst vs. light | Sex: male | 41.8% | 52.7% | 0.2991 |
| | | Abst vs. moderate | Sex: male | 41.8% | 56.2% | 0.3968 |
| | | Abst vs. moderate | Smoking: never | 52.1% | 37.0% | 0.3060 |
| | | Abst vs. moderate | Smoking: current | 21.6% | 33.2% | 0.2536 |
| | | Abst vs. heavy | Sex: male | 41.8% | 59.8% | 0.4936 |
| | | Abst vs. heavy | Smoking: never | 52.1% | 33.6% | 0.3749 |

PREPRINT

| | | | | | | |
|---|---|---|---|---|---|---|
| | | Abst vs. heavy | Smoking: current | 21.6% | 40.8% | 0.4197 |
| | | Abst vs. heavy | Excitement-seeking | 21.7 | 23.0 | 0.2792 |
| | Wine | Abst vs. heavy | Smoking: never | 49.4% | 34.5% | 0.3002 |

Abbreviations: abst. = abstinent

**Supplemental Table S6.2**. Fully adjusted models comparing abstinent participants to each drinking level, and binge-drinkers to non-binge-drinkers, in age- and sex stratified groups

| | | N | β | 95% CI | p |
|---|---|---|---|---|---|
| Occasional (0-2.5 g/day) vs. abstinent | Full sample | 16,838 | 0.018 | -0.006 – 0.043 | 0.1432 |
| | Beer | 2,369 | 0.036 | -0.015 – 0.087 | 0.1632 |
| | Wine | 11,104 | 0.022 | -0.002 – 0.047 | 0.0771 |
| Light (2.5-14.9 g/day) vs. abstinent | Full sample | 33,827 | 0.056 | 0.033 – 0.078 | <0.0001 |
| | Beer * | 5,637 | 0.037 | -0.010 – 0.085 | 0.1248 |
| | Wine | 22,399 | 0.065 | 0.043 – 0.087 | <0.0001 |
| Moderate (15-29.9 g/day) vs. abstinent | Full sample | 8,955 | 0.111 | 0.079 – 0.143 | <0.0001 |
| | Beer * | 2,194 | 0.134 | 0.061 – 0.207 | 0.0003 |
| | Wine | 5,010 | 0.119 | 0.080 – 0.157 | <0.0001 |
| Heavy (30-60 g/day) vs. abstinent | Full sample | 1,839 | 0.075 | -0.009 – 0.158 | 0.0791 |
| | Beer * | 793 | 0.051 | -0.070 – 0.173 | 0.4062 |
| | Wine * | 626 | 0.037 | -0.074 – 0.147 | 0.5151 |
| Binge-drinking yes vs no | Full sample | 8,380 | 0.032 | -0.002 – 0.066 | 0.0654 |
| | Beer | 3,563 | 0.027 | -0.026 – 0.080 | 0.3134 |
| | Wine | 2,958 | 0.055 | 0.001 – 0.110 | 0.0470 |

* Residual non-adjustable imbalance (ASMD>0.25) remained after propensity score weighing, rendering this between-group comparison prone to residual confounding.

PREPRINT
